# Evaluating the direct effects of childhood adiposity on adult systemic-metabolism: A multivariable Mendelian randomization analysis

**DOI:** 10.1101/2020.08.25.20181412

**Authors:** Tom G Richardson, Juha Mykkänen, Katja Pahkala, Mika Ala-Korpela, Joshua A Bell, Kurt Taylor, Jorma Viikari, Terho Lehtimäki, Olli Raitakari, George Davey Smith

## Abstract

**Background:** Individuals who are obese in childhood have an elevated risk of cardiometabolic disease in adulthood. However, whether childhood adiposity directly impacts intermediate markers of this risk, independent of adult adiposity, is unclear.

**Methods and Results:** We conducted a multivariable Mendelian randomization (MR) study to simultaneously evaluate the effects of childhood and adulthood body size on over 100 systemic molecular biomarkers representing multiple metabolic pathways. We first validated UK Biobank-derived genetic risk scores using data on body mass index (BMI) measured during childhood (n=2,427, age: 3-18 years) and adulthood (n= 1,762, age: 34-49 years) from the Young Finns Study (YFS). Results indicated that the childhood score is a stronger predictor of childhood BMI (0.74 vs 0.62 area under the curve (AUC) for the childhood and adult scores respectively), whereas the adult score was a stronger predictor of adulthood BMI (0.57 vs 0.62 AUC). Two-sample MR analyses in a univariable setting using summary genome-wide association study (GWAS) data in up to 24,925 adults provided evidence of an effect of childhood body size on 42 of the 123 metabolic markers assessed (based on P<4.07×10^-04^). Undertaking multivariable MR analyses suggested that the effects for the majority of these metabolic biomarkers (35/42) substantially attenuated when accounting for adult body size. In further analyses, the biomarkers with the strongest evidence of mediating a long-term effect of adiposity on coronary artery disease (CAD) risk were those related to triglyceride-rich very-low-density lipoprotein particles. In contrast, the biomarkers which showed the strongest evidence of being directly influenced by childhood body size (amino acids leucine, isoleucine and tyrosine) provided little evidence that they mediate this effect on adult disease risk.

**Conclusions:** The effects of childhood adiposity on the majority of biomarkers investigated in this study were greatly attenuated when accounting for adult body size. This suggests that the detrimental impact of genetically predicted childhood adiposity on systemic metabolism, as well as subsequent later life risk of CAD, can likely be mitigated through lifestyle modifications during adolescence and early adulthood.

## Introduction

The rising prevalence of childhood obesity contributes greatly to global healthcare burdens (Hruby and Hu, 2015, Viitasalo et al., 2019). Data from the International Childhood Cardiovascular Cohort Consortium suggests that children who are obese who then remain obese as adults have an increased risk of cardiometabolic disease in adulthood (Juonala et al., 2011). In contrast, children with obesity who do not go on to be obese as adults have a risk similar to that of non-obese children. Separating the effects of childhood and adult body size in populations is extremely challenging, however, particularly given that individuals who are overweight during childhood typically remain so as adults (Biro and Wien, 2010, Buscot et al., 2018). Furthermore, as diseases such as coronary artery disease (CAD) are preceded by metabolic dysregulation (Wang et al., 2014, Bell et al., 2018b, Global Burden of Metabolic Risk Factors for Chronic Diseases et al., 2014), and because obesity itself is difficult to reduce (Dombrowski et al., 2014), it is also increasingly important to identify molecular biomarkers responsible for mediating effects of adiposity on disease risk.

We recently demonstrated that the challenge of separating effects of adiposity at different life stages can be addressed using human genetics by applying an approach known as multivariable Mendelian randomization (MR) (Richardson et al., 2020a, Sanderson et al., 2019, Burgess and Thompson, 2015). This method exploits the random assortment of genetic alleles within a population to disentangle the effects of multiple closely related exposures (e.g. body size at different life stages) on disease risk. Moreover, under the principles of MR, these genetic variants are inherited randomly at conception and are thus robust to confounding and reverse causation (Davey Smith and Ebrahim, 2003, Davey Smith and Hemani, 2014).

As illustrated in **Figure 1A**, MR can be applied in a univariable setting to estimate the effects of childhood body size on complex traits and disease outcomes (e.g. a circulating biomarker or CAD). This is referred to as the ‘total effect’ of child body size, which does not account for adult body size in the model. Previously, we identified strong evidence of a total effect of child body size on adult CAD risk (OR: 1.49 per change in child body size category, 95% CI: 1.33 to 1.68) (Richardson et al., 2020a). Multivariable MR allows the effects of child and adult body size to be simultaneously estimated **(Figures 1B & 1C)**, making it possible to estimate the ‘direct effect’ of childhood body size that is not mediated via adult body size **(Figure 1B)**. Similarly, the ‘indirect effect’ can also be estimated which is the contribution mediated along the causal pathway via adult body size **(Figure 1C)**. For example, in the previous analysis on CAD risk, effect estimates from the univariable analysis attenuated to the null when accounting for adult body size (OR: 1.02, 95% CI:0.86 to 1.22), suggesting that child obesity affects CAD only indirectly via adult obesity. Observational associations between childhood obesity and adult CAD may therefore be explained by individuals remaining obese into adulthood. There is strong support from the literature for this indirect effect on CAD (Umer et al., 2017), although fewer studies have investigated the independent effect of child adiposity on intermediate traits measured in adulthood such as circulating biomarkers of systemic metabolism.

**Figure 1:**
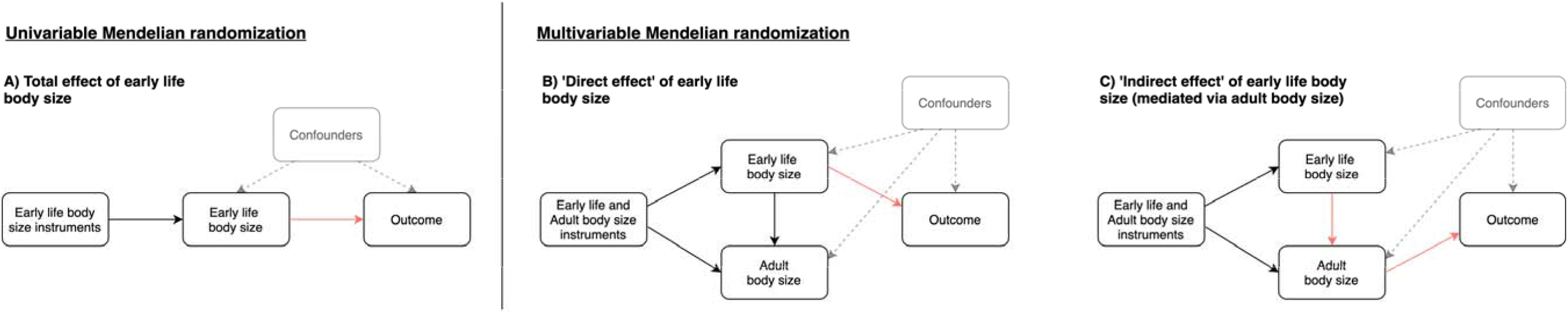
Directed acyclic graphs to illustrate A) univariable Mendelian randomization analyses used in this study to estimate total effects between childhood body size and circulating metabolites B) multivariable Mendelian randomization analyses to estimate direct effects of childhood body size of circulating metabolites and C) applying the same multivariable framework to estimate indirect effects on circulating metabolites mediated along the causal pathway via adult body size.

In this study, we aimed to to comprehensively estimate the direct and indirect effects of childhood body size on detailed biomarkers of systemic metabolism measured via targeted metabolomics in adulthood (Kettunen et al., 2016), and the potential role of these biomarkers in mediating risk for CAD. Firstly, we sought to externally validate our derived genetic scores using data from the Young Finns Study (YFS) to reinforce their capability to separate child and adult body size. Next, in two-sample MR, we estimated the total effect of childhood body size on each of the 123 metabolism-related biomarkers using univariable MR. For metabolic markers with the strongest evidence of a genetically predicted effect in this analysis, we applied multivariable MR to examine evidence of direct or indirect effects after accounting for adult body size. Lastly, we evaluated which biomarkers may potentially mediate the indirect effect of childhood body size on CAD risk, as well as evaluating potential downstream consequences on a wide range of 126 traits and outcomes for markers which may be directly influenced by childhood body size.

## Methods

### Identification of genetic instruments for childhood and adult body size

We previously identified genetic instruments for childhood and adult body size by undertaking a genome-wide association study (GWAS) of 453,169 individuals of European descent from the UK Biobank study (Sudlow et al., 2015). Details have been described previously (Richardson et al., 2020a). In brief, we used questionnaire data asking participants ‘When you were 10 years old, compared to average would you describe yourself as thinner, plumper or about average?’ to define our childhood body size variable. We then harmonized adult measured body mass index (BMI) by converting it into a categorical variable with 3 groups based on the same proportions as the childhood measure (i.e. ‘thinner’, ‘plumper’ and ‘about average’).

GWAS in the UK Biobank study are particularly susceptible to population structure as demonstrated elsewhere (Abdellaoui et al., 2019). We therefore undertook analyses adjusted for age, sex and genotyping chip using the BOLT-LMM software which generates a genetic relationship matrix between samples to account for relatedness and population stratification (Loh et al., 2015). In total, there were 295 and 557 genetic instruments detected for childhood and adult body size respectively based on conventional genomewide corrections (i.e. P<5×10^-08^) **(Supplementary Tables 1 & 2)**. Previous investigations of these instruments in both univariable and multivariable settings with respect to CAD provided strong evidence that they are unlikely to suffer from weak instrument bias based on derived F-statistics **(Supplementary Table 3)**.

We previously attempted to validate these findings by comparing their genetic correlations with results from GWAS of measured childhood obesity (Bradfield et al., 2012) and adult BMI analysed as a continuous variable (Speliotes et al., 2010). Despite the use of recall data in our study to derive the childhood body size instruments, we found that our results were more strongly correlated with measured childhood obesity (r_g_=0.85) compared to adult BMI (r_g_=0.67). In contrast, our adult body size GWAS was very strongly correlated with the adult BMI findings (r_g_=0.96) in comparison to their correlation with childhood obesity (r_g_=0.64). Although these findings suggest that our two sets of genetic instruments can be used to separate the direct and indirect effects of childhood body size using multivariable MR, we set out in this current study to conduct further validation analyses to support this claim.

### Data sources

#### The Cardiovascular Risk in Young Finns Study (YFS)

YFS is a multicenter follow-up study in five cities and their rural surroundings in Finland to evaluate atherosclerotic risk factors from childhood into adulthood. The study began in 1980 when 3,596 participants randomly selected from the national register were examined (boys and girls; ages 3, *6, 9*, 12, 15, and 18 years). Subsequently, follow-up studies have been conducted regularly. Here, BMI data collected at 1980,1983,1986 (from childhood to young adulthood) and 2011 (adulthood) follow-up studies were included. Metabolic traits derived using nuclear magnetic resonance (NMR) spectroscopy were available from the adulthood timepoint in YFS. Local ethics committees approved the study and participants gave written informed consent. Details of the study design have been presented previously (Raitakari et al., 2008). Further details on measures of adiposity, NMR metabolic traits and genotyping for YFS can be found in **Supplementary Note 1**.

#### The Special Turku Coronary Risk Factor Intervention Project (STRIP)

BMI and circulating metabolites data were also available from the STRIP trial at multiple timepoints in early life. STRIP is a prospective randomised life-style intervention trial that began in infancy and continued through childhood and adolescence to early adulthood (Simell et al., 2009). Altogether 1,062 children born in 1989-1991 were recruited at the age of 5 months by the well-baby clinics in Turku, Finland and were randomised into an intervention group (n=540) or a control group (n=522). Here, the STRIP study is treated as a cohort study with analyses conducted with data from participants in both arms.

#### Summary-level data from genome-wide association studies

Summary GWAS data on a total of 123 circulating metabolites from NMR measured in up to 24,925 adults from 14 cohorts (mean age range:23.9 years - 61.3 years) were available from the Kettunen et al (2016) study (Kettunen et al., 2016) (accessible at http://www.computationalmedicine.fi/data). These mostly included lipids (cholesterol, triglycerides) within lipoprotein subclasses, amino acids, fatty acids, glycolysis-related traits including glucose, and inflammatory glycoprotein acetyls. Summary GWAS data on CAD in a sample of 184,305 (60,801 cases and 123,504 controls) were obtained from the Nikpay et al (2015) (Nikpay et al., 2015) study (accessible at http://www.cardiogramplusc4d.org/data-downloads/).

### Statistical analysis

#### Validating genetic instruments in an external cohort of young Finns

We firstly evaluated validation of the genetic scores derived from the UK Biobank study which was particularly warranted given that these instruments are based on self-reported recall data. This was undertaken by investigating the capability of both childhood and adult scores to predict obesity in childhood and adulthood using age- and sex-adjusted logistic regression models in YFS. Age- and sex-specific international BMI percentiles (Cole et al., 2000) were used to extrapolate cutoff points for age 3 to 18 year groups which equate to a BMI of 30 kg/m^2^ in adulthood **(Supplementary Table 4)** (Juonala et al., 2011). Receiver operating characteristic (ROC) curves were generated for these analyses to determine area under the curve (AUC) coefficients. Differences in AUC between age- and sex-adjusted logistic regression models were estimated with the use of the DeLong algorithm (DeLong et al., 1988).

#### Univariable Mendelian randomization

We applied two-sample MR to estimate the total effect of genetically predicted childhood body size on the 123 circulating biomarkers using statistical packages within the MR-Base platform (Hemani et al., 2018)**(Figure 1a)**. This was undertaken using the inverse variance weighted (IVW) method, which uses all SNP-outcome estimates regressed on those for the SNP-exposure associations to provide an overall weighted estimate of the causal effect based on the inverse of the square of the standard error for the SNP-outcome association. We applied a conservative Bonferroni correction (i.e. P<0.05/123=4.07×10^-04^) as a heuristic to allow a manageable number of metabolic biomarkers that are most strongly influenced by genetically predicted childhood body size to be followed-up in this study. However, downstream analyses were also repeated on all 123 biomarkers and are included in supplementary materials for readers interested in investigating these findings based on a less conservative threshold.

We also undertook various sensitivity analyses in this study to improve the robustness of findings. This included applying the MR directionality test (also referred to as the ‘Steiger method’) to support evidence that our genetic instrument influences our exposure before our outcome as opposed to the opposite direction of effect (Hemani et al., 2017). Moreover, we calculated the intercept term for the MR-Egger method for all univariable analyses to indicate whether directional horizontal pleiotropy may be driving results (Bowden et al., 2015).

#### Observational effect estimates and comparison with genetic estimates

Serum NMR-based variables with skewed distributions were log-transformed prior to statistical analyses. A linear regression model was fitted for each variable, with a categorised BMI variable based on the same proportions as those derived in the initial UK Biobank analysis as the explanatory variable and the biomarker measure as the outcome. In YFS, analyses were performed for those who had data on both childhood/young adulthood BMI and adulthood BMI (N=1,508). In STRIP, analyses were performed separately for each age group. Models were adjusted by sex, birth year and age at measurement of the biomarkers in YFS and sex and birth year in STRIP. Genetic estimates derived in the previous analysis were compared with these observational results using a forest plot for visualization.

#### Multivariable Mendelian randomization

Multivariable MR using the IVW method was subsequently applied in a two-sample setting using the Kettunen et al (2016) circulating metabolites GWAS data. This statistical method fits multiple risk factors as exposures (e.g. childhood and adult body size in this study) to simultaneously estimate their genetically predicted effects on an outcome (e.g. a circulating biomarker). This allowed us to estimate the ‘direct’ effect of childhood body size (i.e. the effect after accounting for adult body size) as well as it’s ‘indirect’ effect (i.e. the effect mediated by adult body size) on each metabolic biomarker (as depicted in **Figure 1b** and **Figure 1c)**. We applied this model using all genetic variants for both childhood and adult body size after undertaking linkage disequilibrium (LD) clumping based on r^2^<0.001 to ensure independence of our instruments. Furthermore, we conducted multivariable MREgger analyses to evaluate horizontal pleiotropy for direct and indirect effects (Rees et al., 2017).

#### Evaluating potential downstream consequences on disease outcomes

All circulating biomarkers identified in the initial univariable MR analysis were also further evaluated to determine whether they may mediate the total effect of adiposity on CAD risk. This was undertaken as before using the IVW method and adjusting resulting p-values based on the 123 tests undertaken. For metabolic markers where childhood body size provided evidence of a direct effect based on our multivariable MR analyses based on this conservative threshold, we also evaluated their putative downstream effects in a hypothesis-free manner on 126 diverse traits and disease outcomes curated previously (Richardson et al., 2018) **(Supplementary Table 5)**.

All analyses were undertaken using R (version 3.5.1) and SAS (version 9.4). Forest plots were created using the R package ‘ggplot2’ (Ginestet, 2011).

## Results

### Validation of genetic scores in the Young Finns Study

The validation study in YFS demonstrated that our genetic score for childhood body size is a stronger predictor of childhood obesity compared to our adult body size score (AUCs [95% CI] 0.74 [0.65-0.83] versus 0.62 [0.53-0.72], P=0.02). Conversely, the adult genetic score was a stronger predictor of adulthood obesity based on a conventional threshold of BMI ≥ 30 kg/m^2^(0.62 [0.58-0.65]) compared to the childhood score (0.57 [0.54-0.60], P=0.02). ROC curves illustrating these results can be found in **Figure 2**. These findings therefore support the utility of these genetic instruments to separate the direct and indirect effects of childhood body size, which builds upon the genetic correlation results reported previously **(Supplementary Table 6)**. This separation is likely driven by genetic variants which have a statistically larger or smaller magnitude of effect on body size in the original GWAS compared to adulthood **(Supplementary Table 7)**.

**Figure 2:**
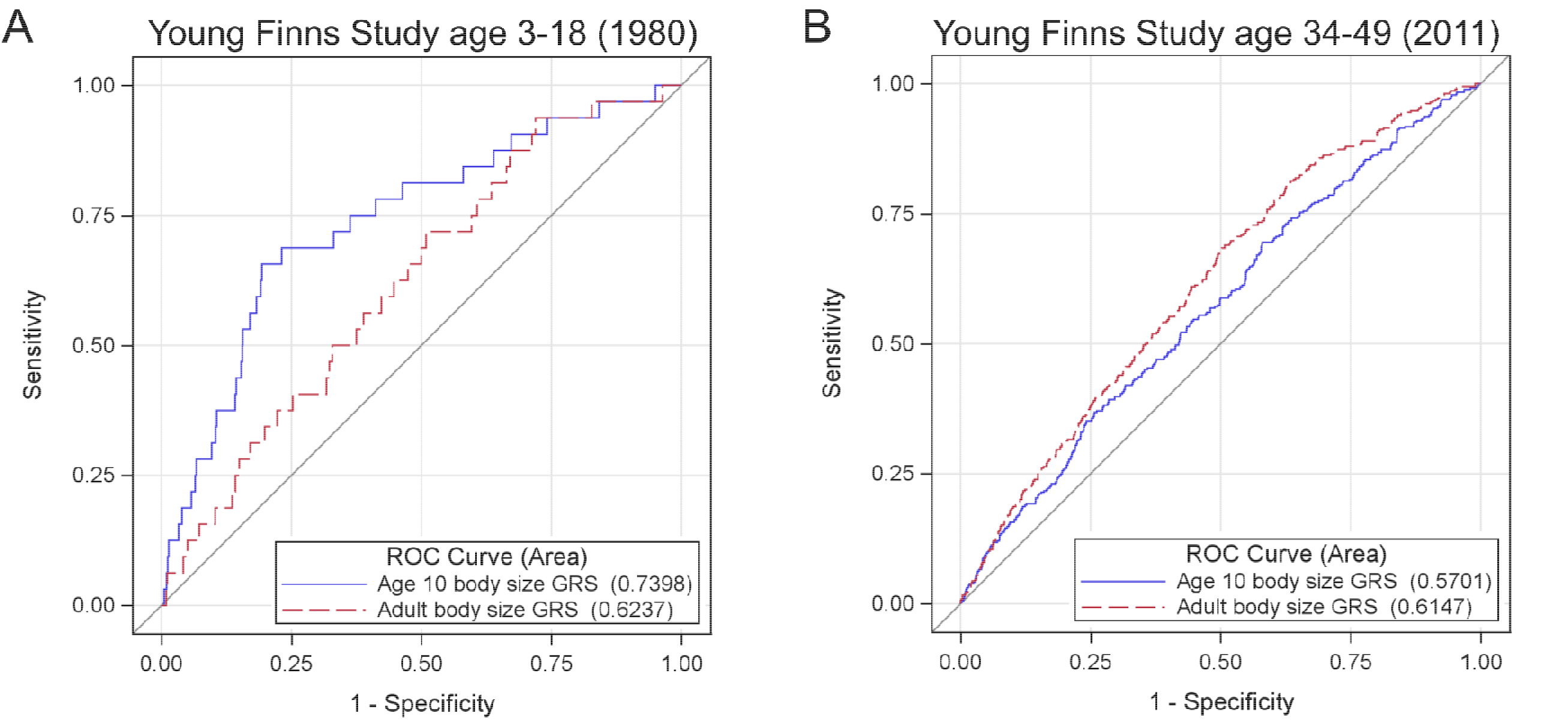
Receiver operator characteristic curves to demonstrate our ability to separate childhood and adult obesity using genetic risk scores. Receiver operator characteristic (ROC) curves to compare the predictive ability of genetic scores for childhood body size (blue) and adult body size (red). A) ROC curve to investigate prediction of adiposity during childhood (N-2,427, age-3-18 years) using cut-offs defined in Supplementary Table 4) and B) ROC curve to investigation prediction of adiposity during adulthood based on BMI≥ 30 kg/m^2^ (N-1,762, age-34-49years).

### Evaluating genetic and observational evidence of a total effect between childhood adiposity and systemic metabolism

Two-sample univariable MR analyses of summary GWAS data provided strong evidence of a total effect between genetically predicted childhood body size and 42 circulating metabolites measured in adulthood (based on P<4.07×10^-04^, **Supplementary Table 8)**. Due to the high correlation which exists between these circulating metabolites, the multiple testing correction applied in this analysis may be overly stringent; estimates for childhood body size on all 123 markers are therefore plotted in **Supplementary Figure 1**. Results suggested that childhood adiposity has an inverse relationship with high-density lipoprotein (HDL) cholesterol-related markers, and a positive relationship with those related to very-low-density lipoprotein (VLDL) cholesterol and triglycerides. There was also strong evidence of a total effect of genetically predicted childhood body size on several amino acids, as well as on glycoprotein acetyls which is a stable marker of cumulative inflammation (P=2.83×10^-08^). Notably, there was very weak evidence for a total effect on glucose (P=0.87). Intercept terms based on the MR-Egger method did not indicate that horizontal pleiotropy was driving these effects **(Supplementary Table 9)** and the MR directionality test supported the direction of effect of childhood body size influencing these circulating biomarkers **(Supplementary Table 10)**.

Observational estimates based on childhood BMI (age 6-12) and circulating metabolites based on analyses undertaken in YFS and STRIP were comparable to those identified from univariable MR analyses as illustrated in **Figure 3 (Supplementary Tables 11 & 12)**. Further analyses at the young adult (age 18-24) and adult (mean age:40.2, range:34-46) time points in YFS and analyses at childhood to adolescence in STRIP suggested that the magnitude of effect for BMI on circulating metabolites typically increased over the life course.

**Figure 3:**
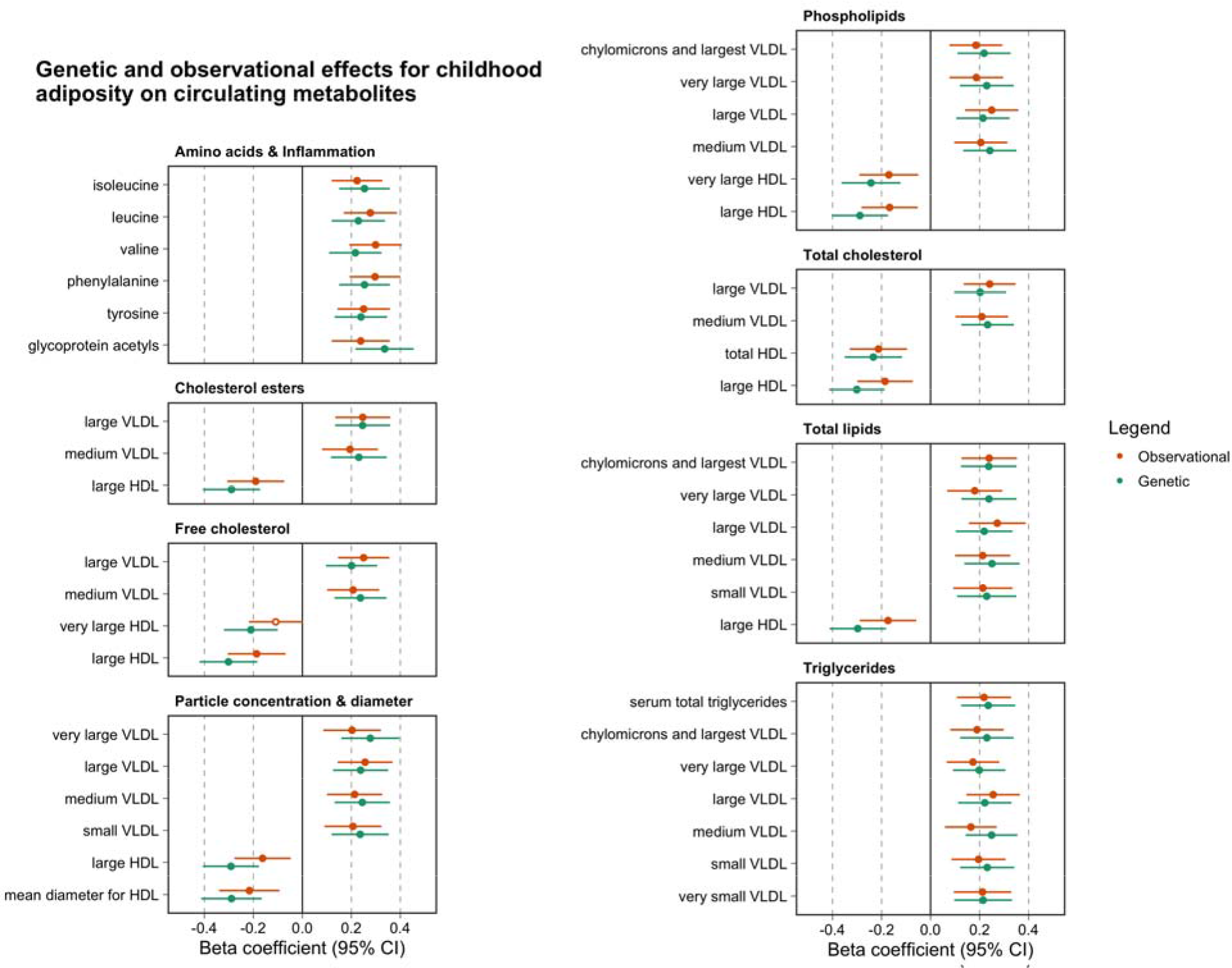
A forest plot depicting observational (orange) and genetic (green) effect estimates between childhood body size (per change in body size category) and circulating metabolites (per standard deviation unit change). Observational estimates were derived using data from the childhood timepoint from the Young Fins Study, whereas genetic estimates are based on two-sample Mendelian randomization analyses using summary data. Note that the observational estimates have been scaled in this figure so that standard errors are the same in both sets of analyses for comparative purposes.

### Using multivariable Mendelian randomization to determine whether childhood adiposity has a direct or indirect effect on circulating metabolites

Applying multivariable MR resulted in the majority of effect estimates identified in the previous analysis (35 out of 42) attenuating to include the null upon adjustment for adult body size **(Supplementary Table 13)**. This suggests that evidence of a total effect between childhood body size and these metabolic biomarkers, as detected in the univariable analysis, is likely attributed to a long-term persistent effect of adiposity across the life course (i.e. not just during childhood). Of the remaining 7 circulating metabolites, the effects of which did not attenuate to the null, there were 3 biomarkers whose central effect estimates for the direct effect of childhood body size were larger in magnitude compared to an indirect effect. These 3 markers were all amino acids; namely leucine (Beta=0.15, SE=0.07, P=0.04), isoleucine (Beta=0.15, SE=0.07, P=0.03) and tyrosine (Beta=0.15, SE=0.07, P=0.03). Effect estimates for childhood and adult body size on these outcomes can be found in **Figure 4A**. Repeating all analyses using the multivariable MR-Egger provided directionally consistent effect estimates to those derived using the IVW method **(Supplementary Table 14)**. We also undertook IVW MVMR analyses on all remaining 123 circulating metabolites in addition to the 42 which survived the heuristic threshold based on Bonferroni corrections **(Supplementary Table 15)**.

**Figure 4:**
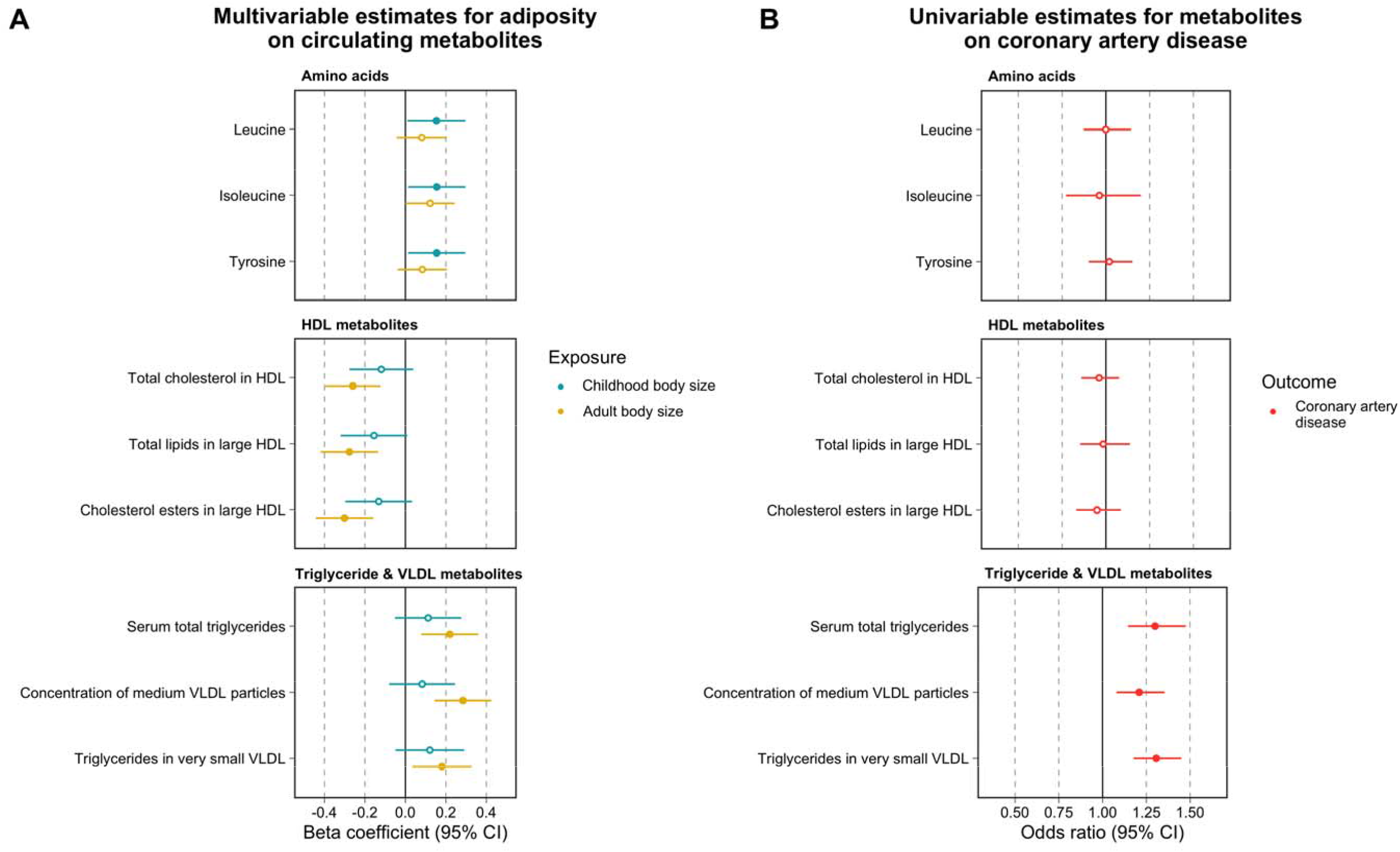
A) forest plot illustrating effect estimates of genetically predicted childhood (blue) and adult (yellow) body size (per change in body size category) on circulating metabolites (per standard deviation change) based on multivariable Mendelian randomization analyses B) A forest plot displaying effect estimates of the same circulating metabolites on risk of coronary artery disease (per standard deviation change) based on univariable Mendelian randomization analyses.

### Assessing the putative mediatory effects of adiposity-influenced metabolic biomarkers on coronary artery disease

Univariable MR analyses provided evidence to suggest that 14 of the metabolic markers identified in the initial analysis influence CAD risk at a level of P<0.05/42=1.19×10^-03^. These were all VLDL-related biomarkers, including serum total triglycerides (OR=1.30, 95% CI=1.17-1.43, P=5.08×10^-05^). As expected, there was a lack of evidence supporting the role of HDL cholesterol-related biomarkers identified in the previous analysis in conferring CAD risk **(Supplementary Table 16)**. We also repeated univariable MR analyses to estimate total effects of all biomarkers (which had at least one genetic instrument based on P<5×10^-08^) on CAD risk, not just the 42 which survived conservative Bonferroni corrections **(Supplementary Table 17)**.

There was no evidence that any of the 3 amino acids which were highlighted in previous analysis altered CAD risk (leucine (OR=1.00, 95% CI=0.86 to 1.13, P=0.99), isoleucine (OR=0.96, 95% CI=0.74 to 1.18, P=0.73) and tyrosine (OR=1.02, 95% CI=0.90 to 1.14, P=0.76)). As before, intercept terms using the MR-Egger method did not suggest directional horizontal pleiotropy was responsible for these results **(Supplementary Table 18)**, nor did the MR directionality test indicate that reverse causality was potentially a major issue for these analyses **(Supplementary Table 19). Figure 4B** illustrates the CAD univariable estimates alongside findings from the previous multivariable analysis for comparison. A subset of circulating metabolites have been grouped into 3 sets within this figure. Firstly, the amino acids which childhood body size had strong evidence of a direct effect on, but very weak evidence that they in turn would influence CAD. Secondly, metabolites related to circulating HDL which, despite providing evidence of being lowered indirectly by childhood body size, are unlikely to have a meaningful effect on CAD (as is becoming increasingly evident (Davey Smith et al., 2020)). Finally, this figure illustrates triglyceride-/ VLDL-related metabolites which evidence suggests play a putative role in mediating the indirect effect of childhood body size on CAD risk.

Lastly, we conducted a hypothesis-free analysis for these 3 amino acids on 126 outcomes **(Supplementary Table 5)** to highlight any potential long-term effects they may mediate between childhood body size and later life disease risk. No results survived multiple testing corrections for tyrosine or leucine (based on the 126 outcomes analyses i.e. P<0.05/126=3.97×10^-04^), including outcomes such as breast cancer and anorexia which may be directly influenced by childhood body size **(Supplementary Table 20)**. We were only able to instrument isoleucine using a single genetic instrument, although there were 10 outcomes which survived multiple testing corrections in this analysis **(Supplementary Table 21)**. However, given that this genetic variant is located at the *GCKR* gene locus, which is known to be highly pleiotropic (Fernandes Silva et al., 2019, Wurtz et al., 2013), we further evaluated the relationship between childhood and adult body size on these 10 outcomes using multivariable MR as undertaken previously. There was weak evidence that childhood body size has a direct effect on these outcomes **(Supplementary Table 22)**, which suggests that it is unlikely that circulating isoleucine mediates any putative effect of childhood adiposity on them.

## Discussion

In this study, we investigated the direct and indirect influence of childhood adiposity on 123 circulating biomarkers of systemic metabolism in adulthood. Based on conservative multiple-testing corrections, there was evidence that genetically predicted body size in childhood has a total effect on 42 of these biomarkers in adulthood. However, accounting for adult body size via multivariable MR suggested that such effects of childhood adiposity are mainly indirect, i.e. they are mediated via adult adiposity. Further analyses suggested that several of these biomarkers related to serum triglycerides and VLDL particles which may putatively mediate the effects of adult adiposity on CAD risk. In contrast, there were 3 amino acids (leucine, isoleucine and tyrosine) which were the only metabolic biomarkers on which childhood adiposity may have a direct effect, although there was no evidence that these amino acids in turn altered CAD risk.

Leveraging data from large-scale GWAS provides a powerful platform to study causal relationships between modifiable risk factors and disease. Conventionally however, MR studies have been limited in their application to temporally segmented effects (Labrecque and Swanson, 2019), which may be attributed in part to the lack of GWAS concerning onset of and subsequent disease progression (Paternoster et al., 2017). As such, interpretation of findings in a univariable setting are confined to genetically predicted exposures based on cross-sectional stages in the life course. Recent methodological developments in MR allow multiple exposures to be investigated in a multivariable framework (Sanderson et al., 2019). Determining whether childhood risk factors have a direct influence on adult disease risk requires modeling them whilst accounting for a measure of the same risk factor taken in adulthood.

Our first objective was therefore to validate a set of genetic instruments for childhood and adult body size, which were previously derived using recall data from the UK Biobank study (Richardson et al., 2020a). This was undertaken using measured BMI data from the Young Finns Study (Raitakari et al., 2008), which suggested that the childhood genetic score is a stronger predictor of childhood BMI in this cohort compared to the adult score, whereas the adult score was a stronger predictor of adult BMI. These analyses provide further validation on top of analyses based on genetic correlation analyses and also validation analyses within the Avon Longitudinal Study of Parents and Children (ALSPAC)(Boyd et al., 2019) **(Supplementary Figure 2)**. Taken together, these findings suggest that our genetic instruments can reliably separate childhood and adult body size as distinct exposures.

Studies from the literature have previously reported strong evidence that BMI causally influences circulating metabolic biomarkers measured in young adulthood (Wurtz et al., 2014) and that such effects of BMI are driven by fat stored centrally (Bell et al., 2018a). Results from our univariable analysis of childhood body size further demonstrate this, as there was evidence of a total effect on 42 circulating metabolites based on stringent Bonferroni corrections for multiple testing (Zheng et al., 2018). In our subsequent multivariable analysis, adjusting analyses for adult body size resulted in 35 of these effects attenuating to the null when accounting for adult body size. This suggests that childhood body size indirectly influences levels of these circulating metabolites via adult body size, as was the case for our previous analysis of CAD (Richardson et al., 2020a). Corroborating evidence of this cumulative, sustained effect of adiposity on circulating metabolites was identified using observational data from the YFS and STRIP cohorts. In these analyses we observed that the magnitude of effect for BMI on these circulating metabolites typically increased over the lifespan. Moreover, our results highlight the importance of accounting for adult measures when investigating the effect of early life exposures on later life disease outcome using MR, which is not always conventionally undertaken in the field (Geng et al., 2018, Fang et al., 2019). Even when observational studies do account for adult body size, they risk inducing collider bias into their analyses by conditioning on a potential mediator (Richardson et al., 2019), which multivariable MR has been shown to be more robust to (Sanderson et al., 2019).

Amongst the 42 circulating metabolites identified in the initial univariable MR analysis, there were 14 biomarkers which may putatively mediate the indirect effect of childhood body size on CAD risk. Notably, these 14 biomarkers were all VLDL- and triglyceride-related. VLDL particles produced by the liver are the major carriers of triglycerides in plasma and are positively associated with both obesity and CAD risk (Triglyceride Coronary Disease Genetics et al., 2010, Mittendorfer et al., 2016). Conversely, there was a lack of evidence from downstream analyses that any HDL-related measure identified in initial analyses influence CAD risk. These findings therefore corroborate evidence from various studies and trial outcomes which support HDL cholesterol or apolipoprotein A-I as non-causal for CAD (Voight et al., 2012, Marz et al., 2017, Richardson et al., 2020b, Karjalainen et al., 2020); although it may still be useful for risk prediction (Davey Smith and Phillips, 2020, Holmes and Davey Smith, 2019).

Our multivariable analysis also suggested that only 3 of the 42 circulating metabolites highlighted by our univariable analyses may putatively be influenced directly by childhood body size. These were the amino acids leucine, isoleucine and tyrosine. Each of these biomarkers has been associated with obesity and cardiometabolic health in previous studies (Suzuki et al., 2019, Moran-Ramos et al., 2017, Taylor et al., 2019), although our analysis suggests that childhood body size may directly influence their levels, potentially in addition to adult body size. However, we found no strong evidence in our study to suggest that these direct effects would have downstream consequences on CAD risk in adulthood.

Our study has several limitations that should be taken into account when interpreting the results. The childhood body size instruments used were derived using recall questionnaire data, which is why we have undertaken analyses in the YFS cohort to provide additional evidence of validation. That said, future GWAS of measured childhood adiposity will be preferable to this score once large-scale sample sizes are available, although for the time-being our scores have been derived in a sample size far larger than any study of measured childhood adiposity. We also acknowledge that, although the premise of MR is to use genetic instruments as proxies to mimic variation in modifiable risk factors, genetically predicted body size may not directly equate to weight change due to lifestyle changes such as diet or exercise as discussed in the early MR papers (Ebrahim and Davey Smith, 2008). Furthermore, the 123 circulating metabolites analysed in this study are predominantly based on lipoprotein lipids which leaves scope to expand upon our analyses in the future. Lastly, we have only used metabolic data from one source in this work for MR analyses due to availability of GWAS summary statistics. Therefore, replication of our results is warranted when those data become accessible.

In conclusion, our findings suggest that the influence of early life adiposity on adult systemic metabolism is predominantly due to an indirect pathway via adulthood body size. Atherogenic VLDL particles may further mediate these effects of sustained adult adiposity on adult CAD risk, whereas evidence of a mediatory role was not supported for amino acids or HDL particles. The impact of childhood obesity on adult cardiometabolic disease risk may therefore be mitigated by reducing adult adiposity, or by targeting intermediate traits like triglyceride-rich lipoproteins if such reductions are infeasible.

## Data Availability

All data analysed in this study is publicly available from the cited sources in the manuscript.

## Acknowledgements

We would like to thank the authors of all the GWAS who made their summary statistics available for the benefit of this work. This work was supported by the Integrative Epidemiology Unit which receives funding from the UK Medical Research Council and the University of Bristol (MC_UU_00011/l). GDS conducts research at the NIHR Biomedical Research Centre at the University Hospitals Bristol NHS Foundation Trust and the University of Bristol. The views expressed in this publication are those of the author(s) and not necessarily those of the NHS, the National Institute for Health Research or the Department of Health. TGR is a UKRI Innovation Research Fellow (MR/S003886/1). JAB is supported by the Elizabeth Blackwell Institute for Health Research, University of Bristol and the Wellcome Trust Institutional Strategic Support Fund (204813/Z/16/Z). KT is supported by a British Heart Foundation Doctoral Training Program (FS/17/60/33474).

The Special Turku Coronary Risk Factor Intervention Project study is funded by the Academy of Finland (grants 206374, 294834, 251360, 275595, and 322112), the Juho Vainio Foundation, the Finnish Foundation for Cardiac Research, the Finnish Ministry of Education and Culture, the Finnish Cultural Foundation, the Sigrid Jusélius Foundation, Special Governmental Grants for Health Sciences Research (Turku University Hospital), the Yrjö Jahnsson Foundation, and the Turku University Foundation. The Young Finns Study is funded by the Academy of Finland: grants 286284, 134309 (Eye), 126925, 121584, 124282, 129378 (Salve), 117787 (Gendi), 41071 (Skidi) and 322098 (for TL); the Social Insurance Institution of Finland; Competitive State Research Financing of the Expert Responsibility area of Kuopio, Tampere and Turku University Hospitals (grant X51001); Juho Vainio Foundation; Paavo Nurmi Foundation; Finnish Foundation for Cardiovascular Research; Finnish Cultural Foundation; The Sigrid Juselius Foundation; Tampere Tuberculosis Foundation; Emil Aaltonen Foundation; Yrjö Jahnsson Foundation; Signe and Ane Gyllenberg Foundation; Diabetes Research Foundation of Finnish Diabetes Association; and EU Horizon 2020 (grant 755320 for TAXINOMISIS and grant 848146 To-Aition); and European Research Council (grant 742927 for MULTIEPIGEN project); Tampere University Hospital Supporting Foundation. MAK is funded by a research grant from the Sigrid Juselius Foundation, Finland.

